# A Wearable Infrared Sensor for Detecting Non-ST Segment Elevation Acute Coronary Syndromes

**DOI:** 10.64898/2025.12.24.25342991

**Authors:** Partho P. Sengupta, Ankush D. Jamthikar, Naveena Yanamala, Kameswari Maganti, Jitto Titus, Sanjeev P. Bhavnani, Lori Daniels, William F. Peacock, Shantanu Sengupta, iSENSE-ACS, multicenter study investigators

## Abstract

**Background:** Non-ST-segment elevation acute coronary syndrome (NSTE-ACS) is conventionally diagnosed using electrocardiography and serial blood biomarker measurements. We investigated a non-invasive, bloodless, and electrode-free diagnostic strategy using a wrist-worn infrared spectrophotometric biosensor (Infrasensor).

**Results:** In a prospective multicenter study of 595 patients with suspected NSTE-ACS enrolled across 13 sites in two countries, participants were stratified into five analytical cohorts. With 200 multi-ethnic controls and a leave-one-cohort-out external validation, a machine learning model detected high-grade coronary obstruction with an area under the receiver operating characteristic curve (AUC) of 0.87 (95% CI: 0.84–0.90), 90% specificity, and 84% positive predictive value—surpassing standard risk scores. A secondary model predicted freedom from NSTE-ACS and adverse outcomes over 30 days with an AUC of 0.89 (95% CI: 0.87–0.92), 99% sensitivity, and 96% negative predictive value.

**Conclusion:** The Infrasensor demonstrated high diagnostic accuracy for high-grade coronary obstruction and 30-day adverse outcome prediction, surpassing conventional risk scores across a prospective, multi-ethnic, multicenter cohort. These findings support its potential as a rapid, non-invasive point-of-care tool for early NSTE-ACS risk stratification.

## Introduction

Acute myocardial infarction (AMI) remains a major cause of mortality in developed countries, affecting nearly 3 million individuals globally and accounting for over 1 million deaths annually in the United States alone^1^. Among the two major subtypes of AMI—ST-elevation myocardial infarction (STEMI) and non-ST-elevation myocardial infarction (NSTEMI)—the proportion of NSTEMI cases has been steadily increasing. This shift has contributed to a rise in non-ST-elevation acute coronary syndrome (NSTE-ACS), which now accounts for approximately 70% of all acute coronary syndrome (ACS) hospitalizations in the United States^2^. Importantly, NSTE-ACS more commonly affects older patients with a greater burden of comorbidities compared to those presenting with STEMI^3^. Moreover, there is a growing consensus that NSTEMI is associated with significantly higher one-year mortality rates than STEMI^4–7^. Furthermore, these studies also highlight that NSTEMI patients tend to receive less optimal treatment, including reduced rates of percutaneous coronary intervention (PCI) and lower adherence to guideline-recommended medications at the time of hospital discharge.

Although diagnostic tools such as cardiac imaging and serum biomarkers have significantly improved, early identification and risk assessment in NSTE-ACS remain complex. The absence of definitive electrocardiographic changes, unlike in STEMI, often hinders prompt diagnosis and appropriate triage. The current standard of care relies on serial high-sensitivity cardiac troponin measurements and clinical risk scores (e.g., HEART), a process endorsed by the 2022 American College of Cardiology Expert Consensus Decision Pathway^8^, which can take several hours to confirm or exclude high-grade obstructive NSTE-ACS. Indeed, traditional rule-out protocols may take 6–12 hours, prolonging emergency department stays and delaying definitive care. This delay is critical because a substantial subset of NSTEMI patients harbors an acute, totally occluded culprit artery, a condition referred to as occlusion MI (OMI), seen in approximately 20–35% of NSTEMI patients who are at high risk for delayed recognition and treatment^9,10^.

Upwards of 60% of OMI cases in NSTEMI are missed when using standard ECG myocardial infarction criteria.^11^ Moreover, such missed occlusions are associated with significantly higher mortality. In one large NSTE-ACS cohort, patients with OMI had about twice as much heart muscle damage and a 72% higher risk of death at 6 months compared with those without occlusion^12^. Additional studies further demonstrate that missed occlusions contribute to adverse outcomes and underscore the limitations of current diagnostic criteria^11,12^. Recent expert commentary highlights the need to reclassify infarction beyond the traditional STEMI/NSTEMI framework^13^. The seriousness of the issue is further compounded since silent or atypical myocardial infarctions constitute a substantial fraction of all events, with population studies estimating 22–40% of MIs occur silently with a high prevalence in diabetic patients^14,15^, underscoring the need for more sensitive, adjunctive diagnostic tools.

Guidelines recommend an “early invasive” strategy defined as coronary revascularization within 24 hours for high-risk NSTE-ACS patients and a delayed invasive approach (within 72 hours) for others^16^, but in practice, access to cardiology services and variable pathways lead to inconsistent times to intervention. Furthermore, every hour of delay in reperfusion for an occluded artery significantly worsens outcomes by increasing infarct size and prolonging myocardial ischemia^17^. Therefore, there is intense interest in novel point-of-care diagnostics that can rapidly stratify NSTE-ACS patients.

Infrasensor^18^ is an artificial intelligence-enabled digital health technology and a wearable, wrist-mounted transdermal infrared spectroscopy device that measures molecular infrared absorption through the skin to detect optical signatures in the epidermis that reflect real-time physiological changes during acute coronary syndromes (Figure 1). It delivers results in under five minutes at the bedside, without the need for venipuncture, specialized equipment, or advanced technical expertise. Translational and early clinical feasibility studies^18–20^ suggest that transdermally derived infrared signals can capture biomarker-associated spectral patterns relevant to ACS, supporting its potential role as a rapid, point-of-care OMI identification tool for patients presenting with suspected ACS. In this study, we evaluated the diagnostic performance of the Infrasensor biosensor against standard ECG and blood-based biomarker assessment for early identification and triage of NSTE-ACS. The primary ‘rule-in’ objective was to detect the presence of severe obstructive coronary artery stenosis, including those identified as OMI via the Infrasensor. The secondary ‘rule-out’ objective was to identify patients who did not have a diagnosis of NSTE-ACS, coronary revascularization, or death within 30 days of admission.

**Figure 1.**
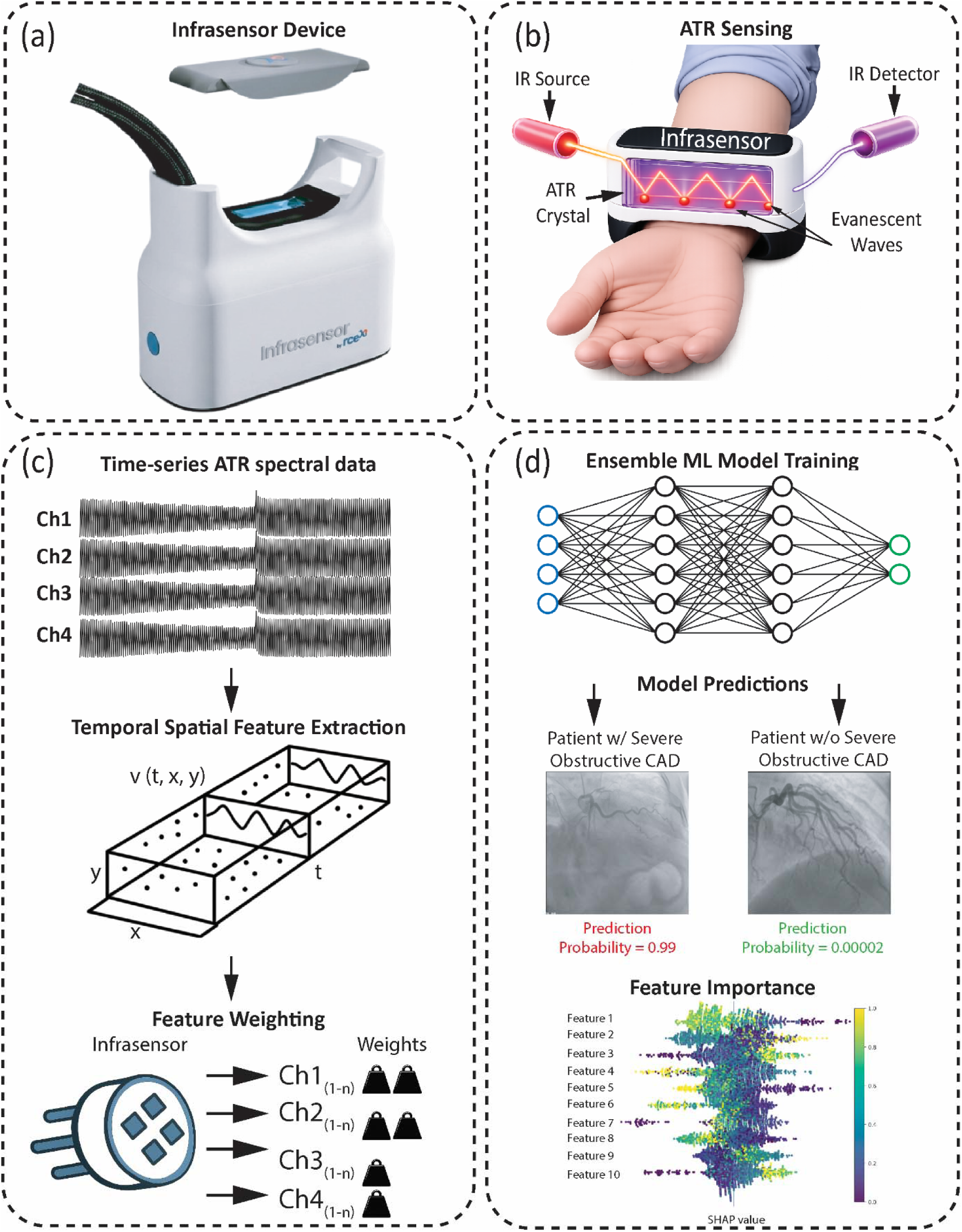
Wrist-wearable infrared sensing system for non-invasive detection of severe obstructive coronary artery disease. (a) The Infrasensor device, a compact wrist-worn unit for continuous ATR spectroscopy. (b) ATR sensing mechanism showing infrared light propagation through the ATR crystal via total internal reflection, generating evanescent waves that penetrate the skin tissue at each reflection point for molecular analysis. (c) Data processing workflow: time-series ATR spectral data from multiple channels are transformed into temporal-spatial features v(t,x,y), followed by feature weighting where channels Ch1□□□□□ through Ch4□□□□□ receive differential importance weights based on predictive value. (d) Ensemble machine learning model architecture processes weighted features to classify patients with versus without severe obstructive CAD, with prediction probabilities validated against coronary angiography. SHAP feature importance analysis identifies the key spectral features driving model predictions.

## Methods

### Trial Design

The iSENSE-ACS trial was a prospective, cross-sectional, multicenter study evaluating the performance of the Infrasensor for the detection of severe coronary stenosis, including OMI in NSTE-ACS. Enrollment began in November 2023 and continued through June 2025. Prior trials have been published that describe the development of the Infrasensor predictor for severe coronary stenosis and OMI in NSTE-ACS. The present study is reported according to the TRIPOD-AI guidelines with a completed checklist provided in the supplementary material. The study was approved by each institution’s IRB and conducted in accordance with Good Clinical Practices. All authors approved the manuscript for publication.

### Study population

The study prospectively enrolled 595 patients with suspected NSTE-ACS across four sites in the United States (n=85) and nine sites in India (n=510). Additionally, 200 healthy individuals were recruited in the United States as controls for model training and calibration. This healthy control cohort consisted of individuals with a very low probability of ACS or obstructive coronary disease and was enrolled to enrich the pool of negatives to support model calibration. These individuals were recruited using a questionnaire to confirm their asymptomatic status and absence of cardiovascular disease.^20^ Each healthy control participant underwent a comprehensive history, clinical examination, ECG, and blood biomarkers (troponin and NT-pro-BNP) to ensure that an acute cardiovascular condition was not present.

Among the study cohort of NSTE-ACS patients, the inclusion and exclusion criteria were designed to be as pragmatic as possible to identify NSTEMI patients and those undergoing coronary angiography. Adult patients aged 18 years or older were eligible for inclusion if they presented to a healthcare facility with symptoms suggestive of ACS and were undergoing clinical evaluation for a diagnosis of NSTEMI. Patients were excluded if they had a diagnosis of STEMI; tattoos, scars, open wounds, or skin lesions at the wrist that could interfere with transdermal signal acquisition or quality; a history of idiopathic pulmonary hypertension; an implantable cardiac device such as a pacemaker, defibrillator, or loop recorder; were pregnant; had an active malignancy; were experiencing hemodynamic or electrical instability, such as cardiogenic shock or unstable atrial or ventricular arrhythmias; had altered consciousness such as delirium; or were unable or unwilling to provide informed consent.

### Primary and Secondary Objectives

The primary ‘rule-in’ objective was to evaluate the performance of a machine-learned Infrasensor predictor for the detection of severe obstructive coronary stenosis, including OMI. The primary effectiveness analysis included individuals who underwent coronary angiography (n = 469). A primary safety analysis was performed to determine the composite rate of readmission for recurrent ACS, coronary revascularization, or death within 30 days of incident hospitalization among all enrolled participants evaluated for NSTE-ACS (n=595).

The following analyses were performed to refine the primary objective:

1. Infrasensor performance across demographic subgroups, including in patients with type I non-STEMI, myocardial infarction with non-obstructive coronary arteries (MINOCA, Type II MI), and unstable angina (including ischemia with non-obstructive coronary arteries, INOCA).
2. Net reclassification index for a positive Infrasensor output stratified by HEART score to determine the yield in troponin-dependent and troponin-independent groups and across low to high-risk patients
3. Moderator analysis across age, gender, and risk factors, angiographic and Troponin subgroups.

The secondary ‘rule-out’ objective was to identify patients who, on clinical grounds, were determined not to have NSTE-ACS. The secondary effectiveness analysis included study participants who underwent coronary angiography and were found not to have severe obstructive coronary stenosis or OMI. The same safety analysis was performed to determine the composite rate of readmission for recurrent ACS, coronary revascularization, or death within 30 days.

Based on geographic location (the United States or India), non-ST-elevation ACS patients were grouped into five analytical cohorts. These groups were formed based on geographic location, aiming for equal cohort sizes to facilitate leave-one-cohort-out external geographic validation. In this framework, models were iteratively trained on four cohorts from different geographical locations and evaluated on the remaining cohort, thereby emulating real-world deployment and providing a rigorous assessment of model generalizability^21^. Specifically, training AI models on multiple diverse cohorts introduces heterogeneity and reduces the influence of hospital-specific patterns, thereby producing models that are more generalizable across different clinical settings^22,23^. An overview of data flow, cohort allocation, and validation strategy, including the integration of the healthy control cohort, is presented in Figure 2a.

**Figure 2.**
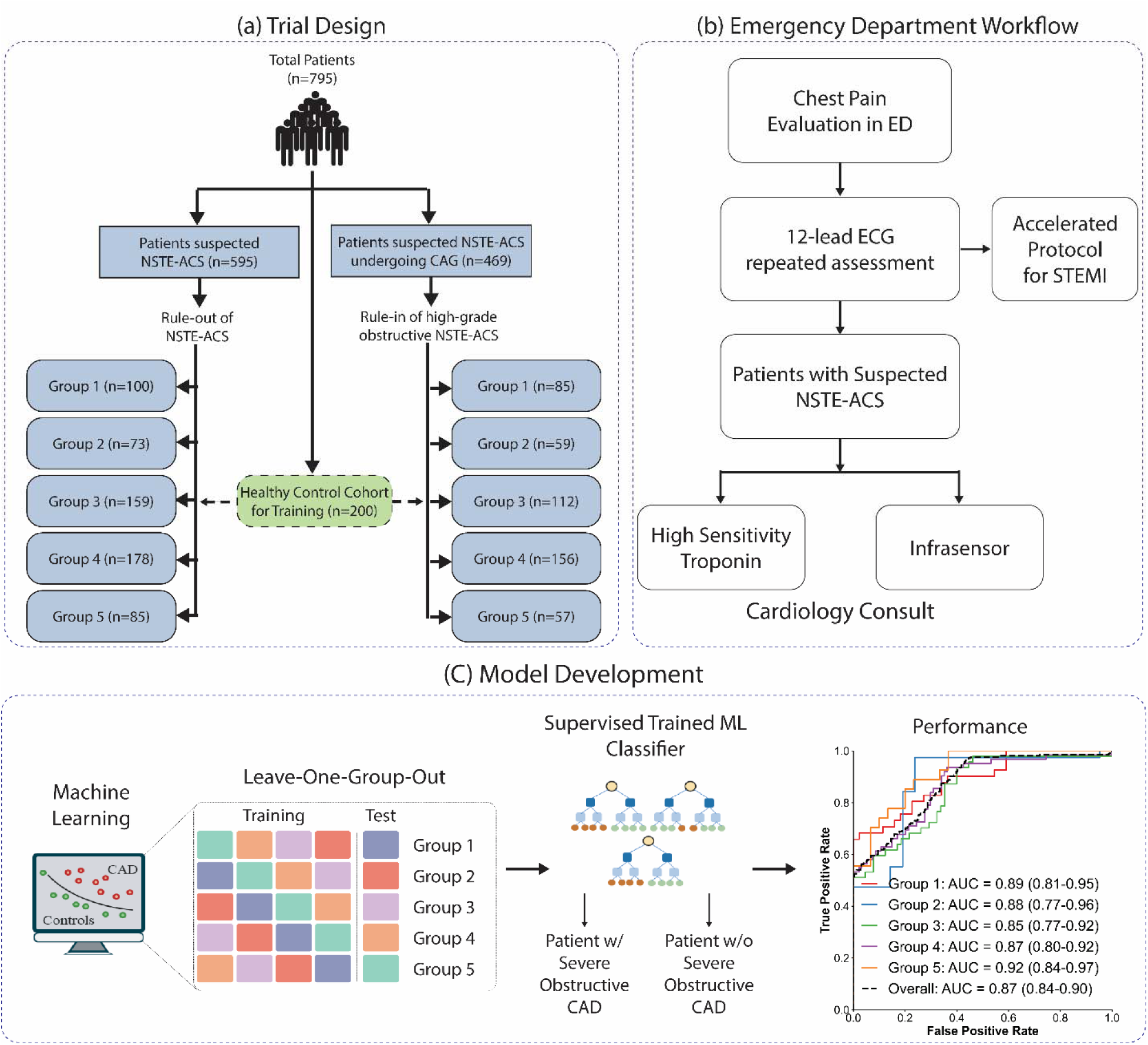
Study design and implementation framework. (a) Trial design and modeling workflow; (b) Infrasensor implementation framework for NSTE-ACS including clinical workflow integration and emergency department workflow integration; (c) Machine learning model development and validation pipeline showing supervised learning approach. CAG = coronary angiography; NSTE-ACS = non-ST-segment elevation acute coronary syndrome

### Study protocol and workflow

All patients received standard of care for ACS from initial presentation through discharge. The study protocol (Figure 2b) included a 12-lead electrocardiogram obtained in the emergency department and repeated as clinically indicated. Serial venous blood samples were collected for high-sensitivity cardiac troponin (hs-cTnI) and N-terminal pro–B-type natriuretic peptide (NT-proBNP) testing. Blood samples were processed locally, with parallel aliquots sent to a central core laboratory for adjudication to ensure consistency across sites. Infrasensor was applied with data collected at any time after an ECG was performed, and before coronary angiography. The reason to perform an Infrasensor test after the ECG was not to introduce a delay in the diagnosis or evaluation of patients with STEMI.

Initial medical treatment and additional diagnostic tests, including echocardiography, were performed as clinically indicated. An invasive coronary angiogram was performed according to guidelines and clinical practice at the individual invasive center. Procedural diagnostic methods, procedural medication, and coronary revascularization were performed at the discretion of the interventional cardiologist. Clinical outcomes were systematically assessed at 30 days through structured phone calls and electronic medical record queries to capture recurrent ACS events, revascularization procedures, or mortality.

### Study data

#### Biomarkers and Clinical Risk Scores

The central lab used the Architect i-STAT High Sensitive Troponin-I (hscTnI) ^24^ (Abbott Laboratories, Abbott Park, IL, USA) assay for all Indian sites, as well as for healthy normals from the US sites. The assay has a recommended sex-specific 99th percentile cut-off point of 15.6□ng/l for females and 34.2□ng/l for males. In the US cohort, the assay varied across sites; the UCSD site used the Elecsys Troponin T Gen 5 assay with a lower limit of detection of 6 ng/L. The 99th percentile is 22 ng/L for males and 14 ng/L for females. The Ben Taub site used a Beckman DXI 600 analyzer to measure hscTnI, with 100 ng/L considered as the threshold for elevation. Rutgers used the Siemens Atellica hscTnI assay with 99^th^ percentile thresholds of 53.53 ng/L and 38.64 ng/L for males and females, respectively. As different assays were used, hscTnI was stratified into normal limit, 1-3 times normal limit, and >3 times normal limit, as categorized in the HEART score. Herein, for consistency, these categories will be referenced as normal, mildly elevated, and elevated hscTnI.

### HEART Score

The HEART score, a validated clinical risk score for acute chest pain, was calculated for each patient. This score incorporates five elements: clinical **H**istory (0-2 points), **E**lectrocardiogram findings (0-2 points), **A**ge (0-2 points), **R**isk factors (0-2 points), and **T**roponin levels (0-2 points), with total scores ranging from 0-10. Higher HEART scores have been shown to correlate with a greater likelihood of major adverse cardiac events (MACE) within 30 days^25^. The HEART score was used in this study for risk stratification and to benchmark the performance of our prediction model.

### Coronary angiography

At each study site, stenosis was visually assessed and reported. Centrally, each site angiogram report was entered into the database as stenosis in the right coronary artery, left anterior descending artery, and left coronary circumflex artery, or left main artery with a maximum level of stenosis categorized as: (1) normal (0%), (2) non-significant/mild or minor (1% to 49%), (3) moderate (50% to 69%), (4) significant/severe (70% to 99%), or (5) total occlusion (100%). For the left main artery, the maximum level of stenosis was categorized as: (1) normal (0%), (2) non-significant/mild or minor (1% to 49%), (3) significant (50% to 69%), (4) severe (70% to 99%) or (4) total occlusion (100%). The severity of each stenosis was further assessed using a CASS-50 angiographic score since this score has a strong angiographic correlation with plaque burden.^26^ All angiograms were adjudicated for the severity of coronary stenosis by the treating interventional cardiologist. To confirm the accuracy of the recorded data, one core lab interventional cardiologist first reviewed all the coronary angiograms for visual assessment of stenosis, who was blinded to local site adjudication, clinical data, and Infrasensor output. All patients with severe obstructive stenosis were also assessed for the TIMI flow grades^27^ and collected according to standard definitions of perfusion: (0) no perfusion, (1) penetration without perfusion, (2) partial perfusion, and (3) complete perfusion.

### Definition of Occlusion MI (OMI)

Occlusion MI was defined as an acutely, totally occluded (100%) coronary artery or a severe (70% to 99%) coronary stenosis with TIMI 0–2 flow^28^ or a High Sensitive Troponin-I assay >200 times the upper limit of normal^29^. All coronary angiograms were first reviewed visually by a blinded interventional cardiologist at the central core laboratory. Cases with significant or severe stenosis (70–99%) or OMI underwent an additional review by a second blinded interventional cardiologist. In instances where discrepancies arose between the local site interpretation and the anonymous core reviews, final lesion severity was determined by majority consensus across the three assessments (local interpretation plus two blinded central adjudications).

### Adjudication of non-ST-elevation ACS

All relevant information, including ECGs, clinical assessment, blood results, coronary angiography, and cardiac imaging, up to 30 days, was reviewed to adjudicate the possibility of type 1 or type 2 NSTEMI using the criteria defined by the 4^th^ universal definition of myocardial infarction.^30^ Patients in the high suspicion category, defined as chest pain with regional wall motion abnormalities on imaging, were assigned a diagnosis of unstable angina. We restricted the definition of unstable angina to those with a normal or hscTnI ≤99th percentile of hscTnI^31^.

### Study endpoints

The primary endpoint was to determine the performance of the Infrasensor predictor on the angiographic identification of high-grade obstructive coronary stenosis, including OMI in NSTE-ACS patients (defined as >70% stenosis in any epicardial coronary artery or >50% stenosis in the left main coronary artery) and/or the need for coronary revascularization during the initial hospitalization.

The secondary endpoint was to determine the yield of a positive and negative Infrasensor output for the detection of high-grade obstructive coronary stenosis, including OMI in NSTE-ACS patients on the composite outcome comprising any of the following: a final discharge diagnosis of NSTE-ACS; coronary revascularization (percutaneous coronary intervention or coronary artery bypass grafting); recurrent NSTE-ACS; or all-cause mortality, assessed within 30 days of admission.

### Advanced Imaging Analysis

#### Focus Cardiac Ultrasound (FCU)

FCU images obtained at participating sites were transferred to a centralized core imaging laboratory for uniform analysis. An expert cardiologist, blinded in the same fashion as above, reviewed all studies using a standardized semi-qualitative approach based on established guidelines from the American Society of Echocardiography. Regional myocardial function was assessed visually using a 16-segment model of the left ventricle. Global left ventricular size and systolic function were also estimated visually and categorized as normal or abnormal. Image quality was assessed and classified as either good or poor. Good-quality studies were defined as those in which at least 50% of left ventricular segments were visualized throughout the cardiac cycle from apical views, allowing reliable visual assessment of wall motion and left ventricular function. Presence of any complications (ventricular septal defect, papillary muscle dysfunction, severe mitral regurgitation, left ventricular aneurysm or pseudoaneurysm, or clot and pericardial effusion was also recorded.

#### CMR Imaging

A subset of patients (n=127) underwent CMR on a Siemens Spectra scanner (Siemens AG, Erlangen, Germany) at a median interval of 2 days to ensure the adequacy of the adjudicated endpoints. Electrocardiographic gated Steady State Free Precession images were obtained in 2, 3, 4, and short axis stack covering the left ventricle from base to apex during repeated breath holds averaging 5 to 10 seconds. T2 STIR imaging, pre-contrast native T1 (ShMOLLI sequence), and T2 maps were obtained in basal, mid, and apical short-axis views. Resting first-pass perfusion imaging using gradient recalled echo sequence in the same three short axis views, along with 1-2 long axis views, was obtained based on the heart rate immediately after injection of 0.125 mmol/kg of gadolinium diethyltriaminepentaacetic acid bolus at 4 ml/sec. Early gadolinium-enhanced imaging was performed 3 min post-contrast in 3 short-axis views (3-, 4-, and 2-chamber) and three long-axis views. Delayed gadolinium enhancement imaging was acquired 8-12 minutes post-contrast using a phase-sensitive inversion recovery sequence after adjusting the inversion time to ensure adequate myocardial nulling and a maximal contrast between myocardium and cavity in the same views. Post-contrast T1 mapping was performed 15 min after contrast injection in 3 short-axis views to calculate extracellular volume.

Biventricular diastolic and systolic volumes, ejection fraction, myocardial mass, and regional wall motion analysis were quantified. Perfusion image analysis was performed according to the American Heart Association 17-segment model. The perfusion defect was determined by visual assessment after the grayscale and image contrast adjustment. Global perfusion was identified as usual if a perfusion defect was noted in <5% of the myocardial segments. The delayed GE analysis was performed using a 17-segment model by dividing the basal and mid-short-axis slices into six segments and the apical slices into four segments. Each myocardial segment was divided into four layers and scored according to the transmural extent of enhancement (none = 0; 0- 5% = 1; 26-50% = 2; 51 to 75% = 3; and 76 to 100% = 4). Delayed gadolinium enhancement was assessed using the full-width at half-maximum method to calculate the acute infarct size (percentage of the left ventricular mass with delayed GE).

### Infrasensor System Description and Data Acquisition

Infrasensor-derived data were recorded during investigational monitoring procedures. The technology is a wearable, transdermal, infrared-spectroscopy-based device (Infrasensor, rce.ai) used to obtain optical readouts; see Figures 1a and 1b. First, the patient’s volar wrist is scrubbed with a 70% alcohol wipe, and the Infrasensor cycles through an automatic 45-second background with a simple button press. Thereafter, the wearable device is strapped to the wrist, with the optical sensor surface contacting the prepped portion of the wrist for 3 minutes, allowing transdermal signal acquisition (Figure 1b).

A detailed technical description of the system has been previously published.^18,19^ Briefly, at the prism–skin interface, the Infrasensor operates in the attenuated total reflection (ATR) regime. When infrared photons strike from a higher-index internal reflection element at angles above the critical angle, Maxwell’s boundary conditions^32^ admit a non-propagating solution in skin with an imaginary normal wavevector, an evanescent field whose intensity decays exponentially over a depth set by wavelength, incidence angle, and the two media’s refractive indices. Quantum-mechanically, this near field behaves analogously to tunneling. However, no net power propagates normal to the interface, a finite probability amplitude samples molecular vibrational dipoles in the superficial dermis/stratum corneum, if a lossy, absorptive medium (skin) is within the decay length, “frustrated” total internal reflection^33^ transfers energy into those vibrational modes, producing ATR absorption features without bulk transmission.

After the measurement, the device was removed, and the data was wirelessly transmitted to a secure, cloud-based Infraomics platform. This platform applies standardized signal preprocessing and proprietary machine learning algorithms for diagnostic inference. Data transmission and storage complied with institutional data security and privacy standards. Data transmission and storage complied with institutional data security and privacy standards.

### Machine learning model development

Machine-learning model development followed a structured five-step pipeline designed to ensure data quality, robustness, and transparency. First, raw Infrasensor recordings underwent standardized signal-quality checks to exclude noisy segments. Second, a comprehensive set of statistical, time-domain, frequency-domain, and time–frequency features was computed from the curated time-series data, followed by outlier detection and feature preprocessing to remove non-informative variables and stabilize feature distributions (Figure 1c). Third, we benchmarked multiple supervised learning algorithms—including logistic regression, support vector machines, random forests, multilayer perceptrons, single gradient-boosting models (LightGBM and XGBoost), and a stacked ensemble in which LightGBM and XGBoost base learners were trained independently, and their predictions served as inputs to a gradient-boosting meta-learner with controllable weights and complementary hyperparameter configurations (Figure 1d). Hyperparameters for all models were tuned using randomized search with early stopping based on validation performance, and model selection was guided by leave-one-cohort-out cross-validation (Figure 2c) to balance discrimination and generalizability; all analyses were performed in Python version 3.11.5 (Python Software Foundation). Comprehensive details on feature engineering, preprocessing protocols, hyperparameter optimization, final model parameters, and comparative performance metrics are presented in Supplementary Material Section I and Supplementary Table 1. Based on these systematic experiments, the model architecture was implemented as a three-stage ensemble architecture utilizing LightGBM, which integrates feature selection with two independently calibrated LightGBM classifiers whose averaged probability outputs yield robust, well-calibrated predictions for binary OMI classification.

### Statistical analysis

Continuous variables were summarized as medians with interquartile ranges (IQR), and categorical variables were summarized as frequencies with percentages. Comparisons across the five study groups were performed using non-parametric methods: the Kruskal–Wallis test for continuous variables and the Chi-square or Fisher’s exact test, as appropriate, for categorical variables. Diagnostic discrimination was assessed using receiver operating characteristic (ROC) analyses, with the area under the curve (AUC) and corresponding 95% confidence intervals (CI) calculated via bootstrapping. AUCs were compared using the DeLong method. Subgroup differences in AUC performance were evaluated using a Q-statistic–based heterogeneity test, which compares the observed variation in effect sizes across subgroups with the variation expected by chance. The resulting heterogeneity p-value (pHET) quantified the likelihood that subgroup-specific AUCs differed beyond random variation; a pHET <0.05 indicated statistically significant heterogeneity. Using a threshold of 0.5, corresponding to the maximum a posteriori (MAP) decision rule^34^,, the model dichotomized predictions by splitting the probability space at the maximum-entropy point. Sensitivity, specificity, positive predictive value (PPV), and negative predictive value (NPV) were calculated at prespecified thresholds, with statistical significance defined as *p*<0.05 (two-sided). All analyses were performed using standard statistical software.

### Multiplicity and control of Type 1 error

The evaluation of Infrasensor performance across the five cohorts did not involve multiple independent hypothesis tests and therefore did not inflate the type I error rate. If independent hypothesis tests had been conducted across five groups at α=0.05, the family-wise type I error rate would increase to approximately 22.6%; however, this scenario does not apply to the present analytic framework. Because our analytic design did not involve multiple independent hypothesis tests across the five cohorts but instead used internal–external cross-validation with pooled inference and a Q-statistic to assess heterogeneity, no inflation of the Type I error rate occurred, and no multiplicity correction was required.

## Results

After exclusions, 795 participants were included in this study, comprising 595 patients with suspected NSTE-ACS and 200 healthy controls (median age 48 years [interquartile range: 33-56], 42% female). Healthy controls were defined as individuals with normal vital signs and basal metabolic index who answered “no” to all questions on the attached questionnaire^20^, making elevated cTnI levels highly unlikely. This cohort has been previously described in detail.^20^

Clinical characteristics of NSTE-ACS patients are presented in Table 1, stratified by five distinct analytical groups based on geographical proximity. All patients underwent standard diagnostic evaluation, including ECG and troponin testing. Across the five groups, distinct differences were observed between the Indian sites (Groups 1–4) and the US site (Group 5). Patients from the US site were older (median 65 vs. 55–59 years, p<0.0001), had a lower proportion of males (56.7% vs. 65.2–78.0%; p=0.06), and had much higher prevalence of smoking (78.9% vs. ≤23.6%; p<0.0001) and obesity (BMI ≥30: 44.7% vs. 11.2–14.0%; p<0.0001).

**Table 1.**
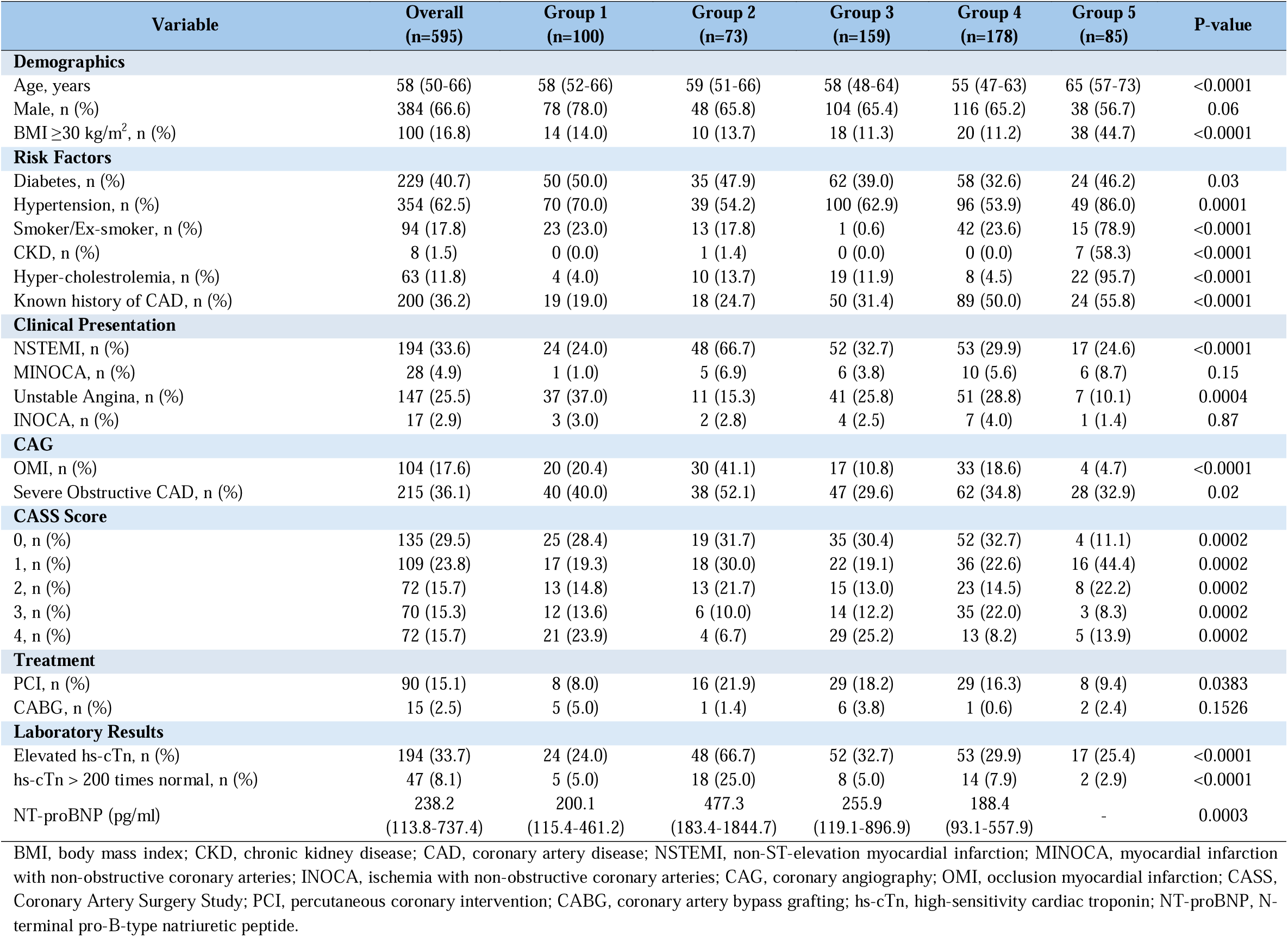
Participant Characteristics and Clinical/Laboratory Measures Across Five Groups.

| Variable | Overall<br>(n=595) | Group 1<br>(n=100) | Group 2<br>(n=73) | Group 3<br>(n=159) | Group 4<br>(n=178) | Group 5<br>(n=85) | P-value |
| --- | --- | --- | --- | --- | --- | --- | --- |
| **Demographics** |  |  |  |  |  |  |  |
| Age, years | 58 (50-66) | 58 (52-66) | 59 (51-66) | 58 (48-64) | 55 (47-63) | 65 (57-73) | <0.0001 |
| Male, n (%) | 384 (66.6) | 78 (78.0) | 48 (65.8) | 104 (65.4) | 116 (65.2) | 38 (56.7) | 0.06 |
| BMI $\geq 30$ kg/m<sup>2</sup>, n (%) | 100 (16.8) | 14 (14.0) | 10 (13.7) | 18 (11.3) | 20 (11.2) | 38 (44.7) | <0.0001 |
| **Risk Factors** |  |  |  |  |  |  |  |
| Diabetes, n (%) | 229 (40.7) | 50 (50.0) | 35 (47.9) | 62 (39.0) | 58 (32.6) | 24 (46.2) | 0.03 |
| Hypertension, n (%) | 354 (62.5) | 70 (70.0) | 39 (54.2) | 100 (62.9) | 96 (53.9) | 49 (86.0) | 0.0001 |
| Smoker/Ex-smoker, n (%) | 94 (17.8) | 23 (23.0) | 13 (17.8) | 1 (0.6) | 42 (23.6) | 15 (78.9) | <0.0001 |
| CKD, n (%) | 8 (1.5) | 0 (0.0) | 1 (1.4) | 0 (0.0) | 0 (0.0) | 7 (58.3) | <0.0001 |
| Hyper-cholesterolemia, n (%) | 63 (11.8) | 4 (4.0) | 10 (13.7) | 19 (11.9) | 8 (4.5) | 22 (95.7) | <0.0001 |
| Known history of CAD, n (%) | 200 (36.2) | 19 (19.0) | 18 (24.7) | 50 (31.4) | 89 (50.0) | 24 (55.8) | <0.0001 |
| **Clinical Presentation** |  |  |  |  |  |  |  |
| NSTEMI, n (%) | 194 (33.6) | 24 (24.0) | 48 (66.7) | 52 (32.7) | 53 (29.9) | 17 (24.6) | <0.0001 |
| MINOCA, n (%) | 28 (4.9) | 1 (1.0) | 5 (6.9) | 6 (3.8) | 10 (5.6) | 6 (8.7) | 0.15 |
| Unstable Angina, n (%) | 147 (25.5) | 37 (37.0) | 11 (15.3) | 41 (25.8) | 51 (28.8) | 7 (10.1) | 0.0004 |
| INOCA, n (%) | 17 (2.9) | 3 (3.0) | 2 (2.8) | 4 (2.5) | 7 (4.0) | 1 (1.4) | 0.87 |
| **CAG** |  |  |  |  |  |  |  |
| OMI, n (%) | 104 (17.6) | 20 (20.4) | 30 (41.1) | 17 (10.8) | 33 (18.6) | 4 (4.7) | <0.0001 |
| Severe Obstructive CAD, n (%) | 215 (36.1) | 40 (40.0) | 38 (52.1) | 47 (29.6) | 62 (34.8) | 28 (32.9) | 0.02 |
| **CASS Score** |  |  |  |  |  |  |  |
| 0, n (%) | 135 (29.5) | 25 (28.4) | 19 (31.7) | 35 (30.4) | 52 (32.7) | 4 (11.1) | 0.0002 |
| 1, n (%) | 109 (23.8) | 17 (19.3) | 18 (30.0) | 22 (19.1) | 36 (22.6) | 16 (44.4) | 0.0002 |
| 2, n (%) | 72 (15.7) | 13 (14.8) | 13 (21.7) | 15 (13.0) | 23 (14.5) | 8 (22.2) | 0.0002 |
| 3, n (%) | 70 (15.3) | 12 (13.6) | 6 (10.0) | 14 (12.2) | 35 (22.0) | 3 (8.3) | 0.0002 |
| 4, n (%) | 72 (15.7) | 21 (23.9) | 4 (6.7) | 29 (25.2) | 13 (8.2) | 5 (13.9) | 0.0002 |
| **Treatment** |  |  |  |  |  |  |  |
| PCI, n (%) | 90 (15.1) | 8 (8.0) | 16 (21.9) | 29 (18.2) | 29 (16.3) | 8 (9.4) | 0.0383 |
| CABG, n (%) | 15 (2.5) | 5 (5.0) | 1 (1.4) | 6 (3.8) | 1 (0.6) | 2 (2.4) | 0.1526 |
| **Laboratory Results** |  |  |  |  |  |  |  |
| Elevated hs-cTn, n (%) | 194 (33.7) | 24 (24.0) | 48 (66.7) | 52 (32.7) | 53 (29.9) | 17 (25.4) | <0.0001 |
| hs-cTn > 200 times normal, n (%) | 47 (8.1) | 5 (5.0) | 18 (25.0) | 8 (5.0) | 14 (7.9) | 2 (2.9) | <0.0001 |
| NT-proBNP (pg/ml) | 238.2<br>(113.8-737.4) | 200.1<br>(115.4-461.2) | 477.3<br>(183.4-1844.7) | 255.9<br>(119.1-896.9) | 188.4<br>(93.1-557.9) | - | 0.0003 |

BMI, body mass index; CKD, chronic kidney disease; CAD, coronary artery disease; NSTEMI, non-ST-elevation myocardial infarction; MINOCA, myocardial infarction with non-obstructive coronary arteries; INOCA, ischemia with non-obstructive coronary arteries; CAG, coronary angiography; OMI, occlusion myocardial infarction; CASS, Coronary Artery Surgery Study; PCI, percutaneous coronary intervention; CABG, coronary artery bypass grafting; hs-cTn, high-sensitivity cardiac troponin; NT-proBNP, N-terminal pro-B-type natriuretic peptide.

Cardiovascular risk factors differed across sites with diabetes was common in all groups (32.6–50.0%; p=0.028), whereas the US site had higher prevalence of hypertension (86.0% vs. 53.9–70.0%; p=0.0001), CKD (58.3% vs. 0–1.4%, p<0.0001), hypercholesterolemia (95.7% vs. 4.0–13.7%, p<0.0001), and previously known CAD (55.8% vs. 19.0–50.0%, p<0.0001).

Regarding the clinical presentation, NSTEMI was the most common (194 patients, 33.6%), ranging from 24.0% to 66.7% across groups (p<0.0001). Unstable angina affected 147 patients (25.5%), varying from 10.1% to 37.0% (p=0.0004). MINOCA (4.9%) and INOCA (2.9%) showed no significant group differences. The Indian cohorts (Groups 1-4, n=510) had higher rates of NSTEMI (34.7% vs 24.6%) and unstable angina (27.5% vs 10.1%) compared to the US cohort (Group 5, n=85).

### Coronary Angiographic Findings

As part of the standard of care at the local sites, 469 out of 595 patients (79%) underwent invasive coronary angiography. Of these 469, 70 patients (14.9%) had triple-vessel disease (CASS 3), while 72 (15.4%) had two-vessel disease (CASS 2), and 109 (23.2%) had single-vessel disease (CASS 1). High-grade obstructive NSTE-ACS, defined as ≥70% stenosis in any epicardial vessel or ≥50% in the left main, was identified in 215 patients (45.8%).

Among the 469 patients undergoing coronary angiography, 91 (19.4%) met criteria for OMI. In the remaining 126 patients who did not undergo coronary angiography, an additional 9 (7.1%) had a highly sensitive troponin I assay >200 times the upper limit of normal, thereby meeting the biochemical criteria for diagnosing OMI. A total of 22 (4.6%) were diagnosed with myocardial infarction with non-obstructive coronary arteries (MINOCA).

A total of 105 NSTE-ACS patients underwent coronary revascularization during same hospitalization: 90 undergoing PCI and 15 undergoing CABG. Additionally, 10 patients were advised PCI and 11 were advised CABG but did not undergo revascularization during hospitalization, while the remaining patients were managed medically. Over a 30-day follow-up period, 11 patients were re-hospitalized with NSTE-ACS, 2 underwent PCI, 3 underwent CABG, and 2 died.

### Diagnostic Performance

#### High-grade obstructive NSTE-ACS

The ROC curve (Figure 3a) demonstrated strong discrimination for the primary endpoint—angiographically confirmed OMI or high-grade obstructive NSTE-ACS—with an AUC of 0.87 (95% CI 0.84–0.90). The Infrasensor showed a specificity of 90% and a PPV of 84% for identifying patients with OMI or high-grade obstructive NSTE-ACS. This indicates that a positive Infrasensor result could effectively and safely guide the need for urgent angiography, with a false-positive rate of 9.8%. Feature importance analysis revealed that spectral and complexity metrics, particularly Shannon entropy and STFT median, were the primary drivers of model predictions (Figure 1d and Supplemental Section II).

**Figure 3.**
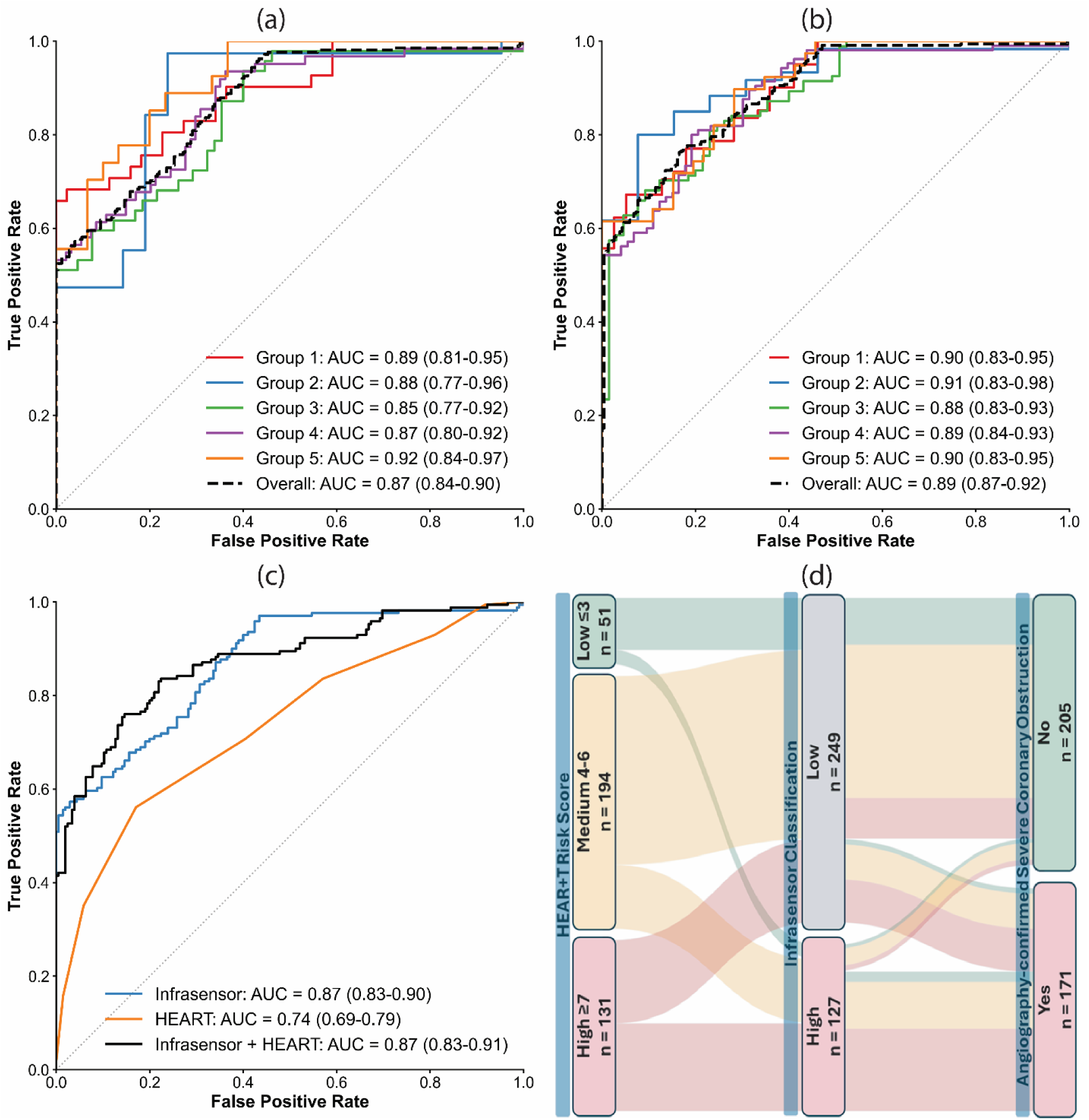
Diagnostic performance and clinical integration of the Infrasensor model. (a) ROC curves for rule-in performance (identifying NSTE-ACS) across individual study groups and combined cohort; (b) ROC curves for rule-out performance (excluding NSTE-ACS) across study groups and combined cohort; (c) Comparative ROC analysis of Infrasensor, HEART score, and combined Infrasensor + HEART model for NSTE-ACS rule-in; (d) Risk reclassification matrix showing Infrasensor refinement of HEART score categories with corresponding rates of angiographic coronary obstruction. AUC = area under the curve; CI = confidence interval; HEART = History, ECG, Age, Risk factors, Troponin.

The proposed Infrasensor-based ML model significantly outperformed the conventional clinical HEART risk score (Figure 3c), which had an AUC of 0.74 (0.69-0.79, p<0.001 vs. Infrasensor). The Sankey diagram (Figure 3d) illustrates the flow of patients from traditional HEART score risk categories to Infrasensor classifications, and ultimately to the presence or absence of high-grade obstructive NSTE-ACS.

Many patients in the medium-risk HEART score category (Supplemental Table 2) were heterogeneously distributed across both rule-in and “observe” arms of the Infrasensor classification. Notably, a substantial proportion (n=49) of patients initially scored as medium risk by HEART were upgraded as ruled-in by the Infrasensor, and a significant subset (n=36) of those were ultimately confirmed to have high-grade obstructive disease. Similarly, several patients in the high HEART score group (n = 64) were reclassified by Infrasensor into the negative category, many of whom (n = 31) did not have high-grade obstructive NSTE-ACS.

Conversely, the Infrasensor “rule-in” arm (n = 67) more precisely identified patients with high-grade CAD, with 63 patients (Supplemental Table 2) having confirmed obstructive disease (Net Reclassification Index of +0.56), enriching the target population for early invasive management. This reclassification led to a clear separation between those with and without disease, as the majority of patients in the rule-in arm were ultimately confirmed to have obstructive CAD. In contrast, those in the “observe” arm were predominantly disease-free.

Subgroup analyses demonstrated no significant effect modification by demographic or comorbidity characteristics, indicating robust and consistent performance of the Infrasensor across clinically relevant subgroups (Table 2).

**Table 2.** Subgroup Analysis of Infrasensor Diagnostic Performance: Pooled AUC With 95% CI and Within-Subgroup Heterogeneity.

| Moderator Variables and Categories | N | AUC (95% CI) | Q (df) | P (het) |
| --- | --- | --- | --- | --- |
| Overall | 469 | 0.87 (0.84-0.90) |  | - |
| **Age (years)** |  |  |  |  |
| <60 | 245 | 0.86 (0.81-0.91) | 0.77 (3) | 0.93 |
| ≥60 | 206 | 0.86 (0.81-0.91) | 2.25 (4) | 0.69 |
| **Sex** |  |  |  |  |
| Female | 146 | 0.90 (0.85-0.94) | 7.44 (4) | 0.11 |
| Male | 305 | 0.85 (0.81-0.89) | 0.18 (4) | 1 |
| **Obese (BMI≥30 kg/cm2)** |  |  |  |  |
| No | 393 | 0.87 (0.84-0.91) | 1.13 (4) | 0.89 |
| Yes | 76 | 0.86 (0.75-0.94) | 1.74 (3) | 0.63 |
| **Diabetes** |  |  |  |  |
| No | 252 | 0.85 (0.79-0.89) | 2.87 (3) | 0.41 |
| Yes | 185 | 0.88 (0.83-0.92) | 4.89 (4) | 0.3 |
| **Hypertension** |  |  |  |  |
| No | 166 | 0.85 (0.78-0.90) | 0.74 (3) | 0.86 |
| Yes | 274 | 0.87 (0.83-0.91) | 2.85 (4) | 0.58 |
| **Troponin (pg/ml)** |  |  |  |  |
| Normal | 282 | 0.86 (0.81-0.91) | 5.61 (3) | 0.13 |
| Mildly Elevated | 92 | 0.88 (0.80-0.95) | 2.53 (3) | 0.47 |
| Elevated | 36 | 0.87 (0.67-1.00) | 4.19 (1) | 0.04 |
| **NT-proBNP (pg/ml)** |  |  |  |  |
| Non-elevated | 96 | 0.85 (0.75-0.93) | 1.34 (2) | 0.51 |
| Elevated | 309 | 0.87 (0.83-0.90) | 0.96 (3) | 0.81 |

BMI, body mass index; NT-proBNP, N-terminal pro-B-type natriuretic peptide; AUC, area under the receiver operating characteristic curve; CI, confidence interval; Q(df), Cochran's Q statistic for heterogeneity with degrees of freedom (df = number of groups - 1); P\_het, P-value for heterogeneity test. P\_het >0.05 indicates no significant heterogeneity in diagnostic performance across study groups.

#### Rule out High-risk ACS

The Infrasensor’s separately trained model on the entire data (n = 595) “rule-out” ACS or related adverse events on a 30-day follow-up with an AUC of 0.89 (Figure 3d, 95% CI 0.87–0.92), demonstrating a robust sensitivity of ∼99% with an NPV of 96%. Importantly, this indicates a rule-out result very rarely misses actual obstruction (∼1–2% miss rate).

### Limited Cardiac Ultrasound SubStudy

We assessed differences in the limited echocardiographic studies performed clinically for patients at the Indian centers (n = 351), divided by predicted phenotypes (Table 3a). The “rule-in” performance of Infrasensor in this subset of patients showed an AUC of 0.88 (95% CI: 0.83-0.92). Across the three groups—severe obstructive coronary artery disease (CAD) (n = 107), remaining NSTE-ACS patients or those with adverse follow-up events (n = 98), and those with non-obstructive or normal coronaries (n = 146)—clinically meaningful differences were observed in LV remodeling and dysfunction, supporting the validity of the adjudication framework used in the study for predicting clinical acute coronary syndrome phenotypes. LV dilation was more common in patients with severe obstructive CAD and in those with composite outcomes compared to those with non-obstructive or normal coronaries (12.7% and 24.2% vs. 2.2%, p < 0.001). Similarly, reduced LV systolic function (LVEF <50%) was observed in 21.3% and 27.5% of the first and second groups, respectively, but in none of the patients with non-obstructive or normal coronary arteries (p < 0.001). Regional wall motion abnormalities followed the same trend, being prevalent in the obstructive and composite-outcome groups (43.8% and 68.9%, respectively) and absent in the non-obstructive group (p < 0.001). Notably, the non-obstructive group exhibited no mechanical complications such as LV aneurysms (5 cases each in the first two groups, none in the non-obstructive group; p = 0.085), papillary muscle rupture (one case in the second group), or LV thrombus (1, 3, and 0 cases across the three groups; p = 0.086).

**Table 3:**
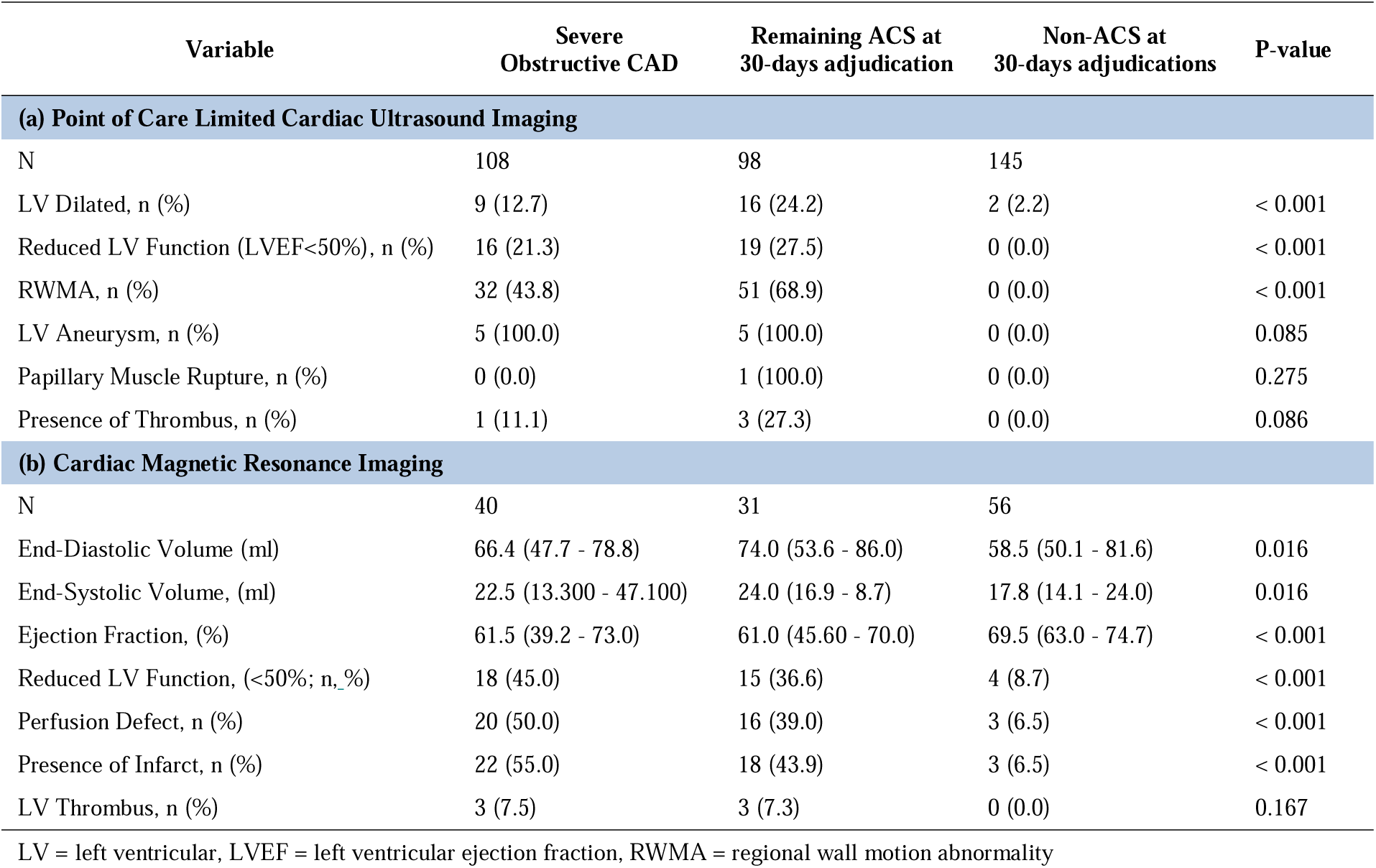
Limited Cardiac Ultrasound and Cardiac Magnetic Resonance Imaging Sub-Analysis in Adjudicated Patient Cohorts.

| Variable | Severe<br>Obstructive CAD | Remaining ACS at<br>30-days adjudication | Non-ACS at<br>30-days adjudications | P-value |
| --- | --- | --- | --- | --- |
| **(a) Point of Care Limited Cardiac Ultrasound Imaging** |  |  |  |  |
| N | 108 | 98 | 145 |  |
| LV Dilated, n (%) | 9 (12.7) | 16 (24.2) | 2 (2.2) | < 0.001 |
| Reduced LV Function (LVEF<50%), n (%) | 16 (21.3) | 19 (27.5) | 0 (0.0) | < 0.001 |
| RWMA, n (%) | 32 (43.8) | 51 (68.9) | 0 (0.0) | < 0.001 |
| LV Aneurysm, n (%) | 5 (100.0) | 5 (100.0) | 0 (0.0) | 0.085 |
| Papillary Muscle Rupture, n (%) | 0 (0.0) | 1 (100.0) | 0 (0.0) | 0.275 |
| Presence of Thrombus, n (%) | 1 (11.1) | 3 (27.3) | 0 (0.0) | 0.086 |
| **(b) Cardiac Magnetic Resonance Imaging** |  |  |  |  |
| N | 40 | 31 | 56 |  |
| End-Diastolic Volume (ml) | 66.4 (47.7 - 78.8) | 74.0 (53.6 - 86.0) | 58.5 (50.1 - 81.6) | 0.016 |
| End-Systolic Volume, (ml) | 22.5 (13.300 - 47.100) | 24.0 (16.9 - 8.7) | 17.8 (14.1 - 24.0) | 0.016 |
| Ejection Fraction, (%) | 61.5 (39.2 - 73.0) | 61.0 (45.60 - 70.0) | 69.5 (63.0 - 74.7) | < 0.001 |
| Reduced LV Function, (<50%; n, %) | 18 (45.0) | 15 (36.6) | 4 (8.7) | < 0.001 |
| Perfusion Defect, n (%) | 20 (50.0) | 16 (39.0) | 3 (6.5) | < 0.001 |
| Presence of Infarct, n (%) | 22 (55.0) | 18 (43.9) | 3 (6.5) | < 0.001 |
| LV Thrombus, n (%) | 3 (7.5) | 3 (7.3) | 0 (0.0) | 0.167 |

LV = left ventricular, LVEF = left ventricular ejection fraction, RWMA = regional wall motion abnormality

### Cardiac Magnetic Resonance Substudy

A subset of patients from the Indian centers (n=127) underwent cardiac magnetic resonance imaging (CMR) to characterize myocardial structure and function and, to additionally investigate the validity of the adjudicated phenotypes predicted by Infrasensor (Table 3b). Across the three groups—severe obstructive CAD (n=40), remaining NSTE-ACS or those with one of the composite follow-up end-points (n=41), and non-obstructive/normal coronaries (n=46)—distinct differences in cardiac structure and function were observed. End-diastolic and end-systolic volumes were higher in the severe obstructive and composite-outcome groups compared with the non-obstructive group (both p=0.016). Ejection fraction was significantly lower in these two groups (median 61.5% and 61.0%) than in patients with non-obstructive coronaries (69.5%, p<0.001). The reduced LV systolic function (LVEF <50%) was correspondingly more prevalent (45.0% and 36.6% vs. 8.7%, p<0.001). Perfusion defects (50.0% and 39.0% vs. 6.5%) and myocardial infarction (55.0% and 43.9% vs. 6.5%) were likewise concentrated in the obstructive and composite-outcome cohorts (both p<0.001). LV thrombus occurred with similar frequency in these two groups and was not observed in patients with non-obstructive or normal coronaries (7.5% and 7.3% vs. 0%; p=0.167). The “rule-in” performance of Infrasensor in this subset of patients was 0.88 (95% CI: 0.80-0.94). Together, these results highlight the Infrasensor’s ability to differentiate meaningful NSTE-ACS cohorts, reinforcing its potential value for early clinical stratification.

## Discussion

This multicenter study demonstrates the clinical utility of a novel, non-invasive, wrist-worn transdermal Infrasensor for the early detection of high-grade obstructive NSTE-ACS, including occlusion myocardial infarction (OMI). Among a diverse, real-world cohort of 795 patients enrolled across 13 sites in India and the United States, the Infrasensor significantly outperformed the current guideline-based risk stratification tool, HEART scores. Importantly, these results were achieved using a transdermal, point-of-care device, offering a rapid, non-invasive alternative to conventional diagnostics. The strength of the study lies in its robust design, incorporating international multicenter enrollment, external validation through a leave-one-cohort-out strategy, and systematic 30-day and clinical follow-up. The CMR substudy confirmed the clinical relevance of this strategy, demonstrating that the Infrasensor identifies patients with larger infarct size, greater prevalence of LV dysfunction, and higher burden of perfusion defects. Collectively, these findings highlight the potential of the Infrasensor to transform early triage, guide invasive management decisions, and improve detection of patients at the highest risk for adverse outcomes in emergency care settings.

To enable early detection of AMI, a wearable sensing device must be able to detect relevant biomolecular changes that occur at the earliest stages of ischemia. In our prior studies, *ex vivo* benchmark experiments demonstrated the feasibility of the Infrasensor device for detecting ischemia-associated biomarkers in the infrared spectral range. These included high-sensitivity troponins, Heart-Type Fatty Acid Binding Protein (H-FABP), creatine kinase-MB, and B-type natriuretic peptide (BNP)^18,35^.

We subsequently validated the device’s performance in clinical settings, demonstrating its ability to detect obstructive coronary artery disease and STEMI^20,35^. In our animal models, ECG changes and lactate elevation occurred within minutes of coronary occlusion, while high-sensitivity troponin levels rose later^36^. Notably, the Infrasensor recorded corresponding spectral signal elevations during this early phase, preceding the detectable troponin rise. These findings suggest that, while the device reliably detects canonical biomarkers such as troponin, BNP, and heat shock proteins (HSPs), it may also be responsive to other biomolecules sharing infrared-active structural features (e.g., amide bonds, carbonyl groups, conformational shifts). The parallel signal rise with early lactate elevation suggests that the Infrasensor may capture a molecular fingerprint of ischemia, encompassing both protein-based markers and early metabolic shifts. While this capability holds promise for *real-time*, preclinical detection of myocardial ischemia, further investigation is needed to isolate and characterize the specific molecular contributors to these early spectral changes.

Our present investigation demonstrates that a rapid, non-invasive infrared spectrophotometric sensor can effectively bridge an important diagnostic gap in acute cardiology. In practice, NSTE-ACS management is fraught with uncertainty; clinicians must identify which patients harbor critical coronary lesions (and thus need immediate intervention) versus those who can be managed conservatively. The Infrasensor’s high accuracy in detecting angiographic-based high-grade obstructive NSTE-ACS has direct clinical implications. By flagging high-risk patients early, even before ECG or blood biomarkers are obtained, the device can expedite definitive treatment. Specifically, patients with OMI who are not promptly recognized often experience reperfusion delays that lead to larger infarcts, heart failure, and higher mortality. In fact, missed occlusions in NSTEMI have been shown to approximately double the risk of death compared to non-occlusive cases^10,37,38^. Implementing the Infrasensor as an adjunct at triage could help ensure that these occult but critical MIs are identified and taken to the cath lab emergently, more in line with STEMI protocols. The present study demonstrates that a positive Infrasensor result is highly predictive of true high-grade lesions (PPV 84%), supporting its use as a reliable trigger for early invasive management. By enabling a targeted strategy, this approach has the potential to shorten time-to-reperfusion in OMI patients, thereby limiting myocardial injury and improving survival. Notably, recent work in redefining ACS around the “Occlusion MI” paradigm emphasizes that timely reperfusion is the key determinant of outcomes, regardless of ST-elevation on the ECG. Our results align with this paradigm by providing a tool for directly detecting occlusion physiology.

Equally important, the Infrasensor can predict freedom from ACS development, future need for PCI, and all-cause mortality. In contemporary practice, the majority of chest pain presentations are ultimately found to be non-cardiac (over 8 million of ∼10 million annual ER chest pain visits in the US). Yet many patients are admitted for prolonged observation and serial troponins out of caution. A negative Infrasensor test, with its 99% sensitivity and high NPV, could allow clinicians to discharge or de-escalate care for patients unlikely to have an ACS or adverse short-term outcome. This can reduce unnecessary hospital admissions, alleviate ED crowding, and allocate resources to patients who genuinely need them. In essence, the device functions as a “throughput optimization tool”, helping to stratify chest pain patients efficiently at the point of care. It is worth noting that the sensor’s output is available within minutes of triage, whereas serial biomarker results take hours. By integrating Infrasensor results with standard clinical assessments (ECG, history, troponin), emergency physicians and cardiologists can make more informed decisions faster. Importantly, a *positive* result would reinforce the urgency of intervention, while a *negative* result would provide reassurance, for instance, in combination with low troponin and a benign ECG. The net effect is a more personalized and timely management of NSTE-ACS: those who need early catheterization receive it without undue delay, and those who do not can avoid invasive testing.

The strong performance of our model suggests that non-invasive biosensors and machine-learning algorithms may soon augment or even partially substitute specific lab tests in ACS diagnosis. Additionally, artificial intelligence applied to subtle ECG features or cardiac ultrasonography has been shown to improve detection of acute occlusion beyond standard criteria^28^. The Infrasensor approach is synergistic with such advances. Together, AI-enhanced ECG analysis and infrared spectral analysis could dramatically improve the early detection of patients with “NSTEMI” who are, in fact, suffering from OMI and need urgent care. Moving forward, it will be valuable to integrate these technologies into clinical workflows and validate their impact on patient-centered outcomes (e.g., infarct size, heart failure rates, mortality). Future prospective trials could test an algorithm that triggers immediate cath lab activation when a positive wearable sensor or AI-ECG is detected, potentially closing the outcome gap between OMI and STEMI patients.

A particularly compelling aspect of the Infrasensor is its potential to improve care in resource-limited environments. Patients in rural or underserved areas often face delayed diagnoses and transfers for acute cardiac care, which contributes to worse outcomes in acute MI. Many smaller hospitals lack on-site cardiology or advanced labs; some may still rely on send-out troponin tests or have no access to urgent angiography. In these scenarios, a portable, easy-to-use device that provides immediate insight into whether a patient’s chest pain is due to a high-grade coronary occlusion can be transformative. For example, an emergency physician in a community hospital could apply the Infrasensor at triage for a chest pain patient with an equivocal ECG. A *positive* result (indicating a likely high-grade obstructive NSTE-ACS) would prompt expediting that patient’s transfer to a tertiary center for possible percutaneous coronary intervention, effectively “flagging” the patient who cannot afford to wait. Conversely, a *negative* result in a low-risk clinical picture might give confidence to manage the patient locally or non-invasively, conserving scarce tertiary resources. By improving early discrimination, the device can help mitigate disparities in care. As Kawa *et al.* note, rural ACS patients suffer higher morbidity in part due to delayed diagnosis and access to intervention^39^. Deploying point-of-care diagnostics like the Infrasensor in ambulances, urgent care clinics, or small regional hospitals could bring advanced cardiac triage capabilities to settings that lack immediate specialist consultation. The device’s design (wrist-worn, battery-operated, cloud-connected) also lends itself to telemedicine integration; for instance, the sensor’s data could be uploaded and interpreted remotely by cardiologists, similar to how remote ECG over-reading is done, enabling specialist input even when none is on site. Such forward-thinking applications could extend acute cardiac care to populations that historically have limited access. Moreover, in global health contexts where advanced imaging or lab infrastructure is scarce, a 5-minute non-invasive test for ischemia would dramatically accelerate the evaluation of chest pain, ensuring that those with true acute coronary occlusions are prioritized for transfer to the nearest facility with revascularization capability.

The findings of this study should be interpreted in the context of several limitations and opportunities for future exploration. First, the study focused exclusively on patients already within hospital-based care pathways. As such, the impact of Infrasensor-guided triage on hard clinical outcomes—such as reduction in myocardial infarction (MI) size or mortality—remains to be determined through randomized controlled trials in both emergency and pre-hospital settings.

Although external validation across geographically and demographically diverse cohorts enhances the generalizability of our findings, successful real-world deployment will require structured clinician training. Ensuring appropriate interpretation and action based on Infrasensor results is critical. While the device showed a low false-positive rate (∼10%), these instances could still lead to unnecessary invasive procedures. Similarly, the false-negative rate (∼6% risk of adverse 30-day outcomes despite a negative test) underscores the importance of using this tool as an adjunct—not a replacement—for clinical judgment and standard diagnostic modalities.

Another key limitation is that the biological content being measured by the Infrasensor remains incompletely understood. While the observed parallel between the sensor’s early signal rise and biomarkers such as lactate and troponin suggests that the device may detect a broader molecular fingerprint of ischemia, additional mechanisms may also be at play. Specifically, biophysical changes in the skin occurring during acute coronary syndromes (ACS)—such as alterations in perfusion, oxygenation, or tissue composition—may influence the epidermal infrared absorption properties that underlie the Infrasensor’s signal. While a detailed molecular and biophysical analysis is still required, the device’s robust performance suggests that these latent changes in infrared absorption associated with ACS can be reliably predicted.

To enhance diagnostic performance, future iterations of the algorithm may integrate patient-specific variables or serial measurements. Additional studies could also assess the utility of the Infrasensor in other clinical scenarios—such as screening stable but high-risk patients in outpatient chest pain clinics, or monitoring patients post-percutaneous coronary intervention (PCI) for recurrent ischemia.

Lastly, a formal cost-effectiveness analysis will be critical to understanding how this technology can be scaled in real-world healthcare systems. While upfront implementation costs must be considered, the potential savings from avoiding missed occlusion myocardial infarctions (OMIs) and reducing unnecessary admissions may be substantial.

## Conclusion

This prospective multicenter study provides evidence that a wearable infrared spectroscopic sensor can rapidly and accurately detect high-grade obstructive NSTE-ACS, outperforming traditional risk stratification. By enabling early identification of “hidden” occlusion myocardial infarctions and safely ruling out low-risk patients, the Infrasensor could usher in a paradigm shift in emergency cardiac care. The integration of this point-of-care device into standard NSTE-ACS pathways has the potential to enhance early triage and improve clinical outcomes, particularly as we strive to deliver timely, equitable care for all patients with acute coronary syndromes.

## Supporting information

Suplemental Material Checklist

## Funding Declaration

This study is sponsored by RCE technologies, Carlsbad, CA, USA

## Ethics Declaration

The study was approved by each institution’s IRB and conducted in accordance with Good Clinical Practices

## Disclosures

PPS is supported by grants from the National Institute of Health/ National Heart, Lung, and Blood Institute (Award 1R01HL173998-01A1, 3U01HL088942-17S, 1P50MD017356-01) and National Science Foundation (Award #2125872). He has served on the Advisory Board of RCE Technologies and HeartSciences and holds stock options; received grants or contracts from RCE Technologies, HeartSciences, Butterfly, and MindMics; and holds patents with Mayo Clinic (US8328724B2), HeartSciences (US11445918B2), and Rutgers Health (62/864,771; US202163152686P; WO2022182603A1; US202163211829P; WO2022266288A1; and US202163212228P).

N.Y. declares grants or contracts from MindMics, RCE Technologies, HeartSciences and Abiomed; receives consulting fees from Turnkey Learning and Turnkey Insights; receives payment or honoraria and support for attending meetings or travel from West Virginia University (WVU) and the National Science Foundation; is an advisor or board member for Research Spark Hub and Magnetic 3D; is an adjunct professor or faculty member at Carnegie Mellon University; is an editorial board member for the American Society of Echocardiography; is a special government employee of the Center for Devices and Radiological Health at the US Food and Drug Association; and holds patents with Rutgers (US202163152686P, WO2022182603A1, US202163211829P, WO2022266288A1, US202163212228P, and WO2022266291A1) and WVU (invention numbers 2021-20 and 2021-047).

W.F.P. holds option equity with RCE Technologies.

J.T. is an employee of RCE Technologies.

S.P.B. is a special government employee and former senior medical officer at the U.S. Food & Drug Administration; has served as a consultant or is on the advisory board of RCE Technologies, Conneqt Health, Samsung Research America; has been on the speaker’s bureau of General Electric Healthcare and Pfizer Patient Advocacy; serves on the scientific advisory board of the Cardiac Science Research Consortium; and is executive director of the Institute for Systems Intelligence and Global Health Transformation (INSIGHT).

All the other authors have nothing to disclose.

## Author Contributions Statement

Design and implementation: PS, JT, SS; Concept and Design: PS, JT, SB, NY, WP, SS; Manuscript Design and editing: JT, AJ, PS, SB, KM.

## Data Availability Statement

The clinical and feature datasets generated and analyzed during the current study are not publicly available due to institutional regulations and the terms of our ethics/IRB approval, which do not permit open sharing of individual-level patient data. De-identified data underlying the main results, together with a data dictionary, are available from the corresponding author upon reasonable request and subject to institutional approvals and a data use agreement.

## Code Availability Statement

Custom code used for data preprocessing, model development, and statistical analysis is available from the corresponding author upon reasonable request for academic research purposes.

## Data Coordinating Center(s)

### INDIA

Shantanu Sengupta – Sengupta Hospital and Research Institute, Nagpur, MS, India

Mahesh Fulwani - Shrikrishna Hrudalaya, Nagpur, MS, India

Aziz Khan - Crescent Hospital, MS, India

Niteen Deshpande - Spandan Heart Institute, Nagpur, MS, India

Pramod Mundra - Platina Heart Institute, Nagpur, MS, India

Uday Mahorkar - Avanti Institute, Nagpur, MS, India

Anuj Sarda - Intima Heart Institute, Nagpur, MS, India

Anil Jawahirani - Central India Cardiology Hospital and Research Institute, Nagpur, MS, India

Sushant Patil - Elixir Metrocity Hospital, Nagpur, MS, India

### USA

Partho P Sengupta – Rutgers Robert Wood Johnson Medical School, New Brunswick, NJ, US

Frank W Peacock – Baylor College of Medicine, Houston, TX, US

Brian Tiffany – Dignity Health, Chandler, AZ, US

Lori B Daniels – University of California San Diego, CA, US

## Acknowledgements

Debasish Bhadra – RCE Technologies, Carlsbad, CA, US

Sabrina Lynch – RCE Technologies, Carlsbad, CA, US

Kunda Mugalmare – Sengupta Hospital and Research Institute, Nagpur, MS, India

Marc Sandhaus – Rutgers Robert Wood Johnson Medical School, New Brunswick, NJ, US

Michael S. Huang – Rutgers Robert Wood Johnson Medical School, New Brunswick, NJ, US

**Correspondence and requests** can be addressed to Partho P. Sengupta, MD, DM, FACC, FASE, Rutgers Robert Wood Johnson Medical School, Division of Cardiovascular Disease and Hypertension, 125 Patterson St, New Brunswick, NJ – 08901.

## Notes

### Clinical Trial

NA

### Author Declarations

IRB of each of the institutions including Rutgers University gave the ethical approval for this work.

### Summary of Updates

This revision was made to correct the study duration in the Methods - Trial Design section. The revised sentence is “Enrollment began in November 2023 and continued through June 2025.”

