## Supplemental Material for "A Wearable Infrared Sensor for Detecting Non-ST Segment Elevation Acute Coronary Syndromes"

**Supplementary Material**

I. **Machine-learning model development**

*Feature engineering and preprocessing*

From each Infrasensor time series, we derived a structured feature set comprising statistical descriptors (mean, median, standard deviation, variance, range, interquartile range, skewness, kurtosis), percentiles (1st, 5th, 10th, 25th, 75th, 90th, 95th and 99th) and median absolute deviation. Time-domain energy and dynamics were summarized using root mean square (RMS), total energy, signal power, linear trend coefficients, mean change, mean absolute change and autocorrelation at lags 1 and 2. Waveform morphology was characterized by zero-crossing rate and peak-to-peak amplitude. Frequency-domain features were obtained from the magnitude spectrum of the fast Fourier transform (mean, standard deviation, minimum and maximum) and from Welch power spectral density estimates (mean, standard deviation, minimum and maximum). Complexity was quantified using Shannon entropy and unique value counts. Time–frequency structure was captured via short-time Fourier transform (STFT) spectrograms, from which mean, standard deviation, minimum and maximum values across time and frequency were extracted. Long-term dependence was summarized using the Hurst exponent. Features with near-zero variance or excessive missingness were excluded; remaining features were imputed (median within training folds) and scaled as needed.

*Candidate models and hyperparameter optimization*

We evaluated multiple classifiers: logistic regression, support vector machines (SVM), random forests, multilayer perceptrons (MLP), LightGBM and XGBoost gradient-boosting models and a stacked ensemble combining LightGBM and XGBoost. Hyperparameters for each model family were tuned using randomized search with nested cross-validation. For LightGBM and XGBoost, the search space included number of estimators, learning rate, maximum tree depth, minimum child samples, subsample ratio and column sampling rate. For tree-based models, class imbalance was addressed via class weights (positive:negative ≈ 5:1). Early stopping rounds were used for boosting models based on validation loss. All model development was implemented in Python 3.11.5 (Python Software Foundation), using scikit-learn, LightGBM and XGBoost libraries.

*Stacked Ensemble Model*

For the stacked ensemble, LightGBM and XGBoost base learners were first trained independently on the full feature set using optimized hyperparameters. Out-of-fold predicted probabilities from each base learner were then used as two-dimensional meta-features to train a gradient-boosting meta-learner, which produced the final prediction. This stacking strategy enabled a dual ensemble with controllable weighting and complementary inductive biases, and was evaluated within a leave-one-cohort-out cross-validation scheme.

*Final three-stage LightGBM ensemble*

The final deployed architecture, described in the main text, consists of: (i) a Stage-1 LightGBM feature-selection model used to rank and select features; (ii) two Stage-2 LightGBM base classifiers with complementary configurations—Model 1 and Model 2; and (iii) Stage-3 ensemble aggregation, in which calibrated probabilities from the two models are averaged to obtain the final probability estimate, with a nominal decision threshold of 0.5.

Supplemental Table 1. List of hyperparameters for model development

| **Stage** | **Model / Component** | **Hyperparameter / Setting** | **Value** |
| --- | --- | --- | --- |
| Stage 1 – Feature selection | LightGBM feature-selection model | n_estimators | 550 |
|  |  | learning_rate | 0.05 |
|  |  | num_leaves | 32 |
|  |  | max_depth | −1 (unlimited) |
|  |  | min_child_samples | 22 |
|  |  | subsample | 0.8 |
|  |  | colsample_bytree | 0.8 |
|  |  | random_state | 41 |
| Stage 2 – Base classifier 1 | LightGBM Classifier (Model 1) | n_estimators | 300 |
|  |  | learning_rate | 0.05 |
|  |  | num_leaves | 32 |
|  |  | max_depth | −1 |
|  |  | min_child_samples | 23 |
|  |  | subsample | 0.8 |
|  |  | colsample_bytree | 0.5 |
|  |  | class_weight | {1: 5, 0: 1} |
|  |  | random_state | 31 |
|  |  | Calibration method | Sigmoid (Platt scaling, CalibratedClassifierCV) |
| Stage 2 – Base classifier 2 | LightGBM Classifier (Model 2) | n_estimators | 620 |
|  |  | learning_rate | 0.04 |
|  |  | num_leaves | 48 |
|  |  | max_depth | −1 |
|  |  | min_child_samples | 20 |
|  |  | subsample | 0.8 |
|  |  | colsample_bytree | 0.6 |
|  |  | class_weight | {1: 5, 0: 1} |
|  |  | random_state | 99 |
|  |  | Calibration method | Sigmoid (Platt scaling, CalibratedClassifierCV) |
| Stage 3 – Ensemble aggregation | Probability averaging | Aggregation formula | (p1 + p2)/2 |
|  |  | Decision threshold | 0.5 |
|  |  | Class distribution handling | Imbalanced, 5:1 weighting for positive class in base models |

II. **Feature Importance Analysis**

Supplemental Figure 1 displays the SHAP beeswarm plot visualizing feature importance and directional impact for the Infrasensor model. Features are ranked by importance (top = most influential), with each point representing a patient. Horizontal SHAP values indicate feature impact: positive values (right of zero) increase predicted probability of obstructive coronary disease, negative values decrease it. Color represents feature magnitude (yellow/green = high, purple = low). Shannon entropy demonstrated highest importance, with elevated values consistently associated with positive SHAP values, indicating greater signal complexity predicts obstruction. Similarly, STFT median showed strong positive associations when high. Conversely, temporal mean absolute change exhibited inverse relationships, with high values reducing obstruction probability. The horizontal spread indicates discriminative contribution—wider distributions (Shannon entropy, STFT median) distinguish obstructive from non-obstructive cases more effectively. Overall, the model integrates complementary features across spectral, temporal, and complexity domains, with spectral and complexity metrics providing the strongest discriminative power.

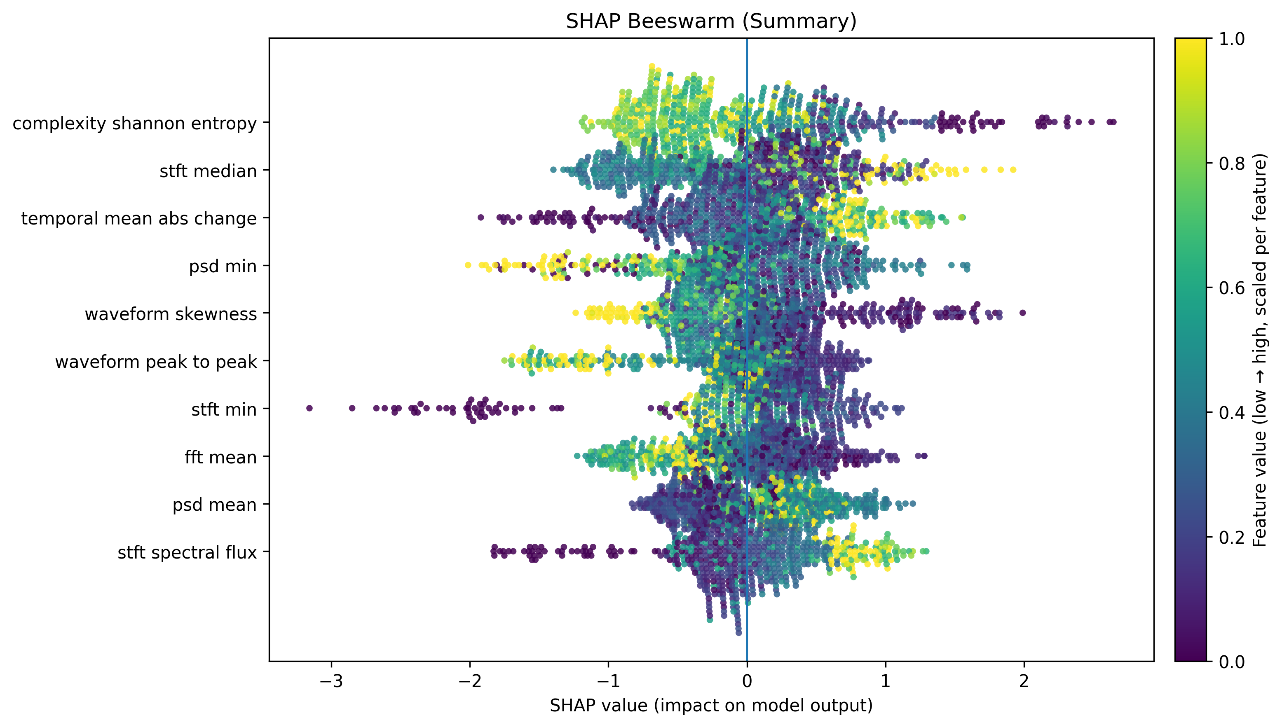

**Supplemental Figure 1.** Global Feature Importance and Directional Impact on Infrasensor Model Predictions Using SHAP Analysis

**Supplemental Table 2. Complete reclassification pathways from HEART score to angiographic outcome for “rule-in” performance of Infrasensor**

| **HEART**  **Score** | **Infrasensor** | **Severe Coronary Obstruction** | **N** | **% of HEART Category** |
| --- | --- | --- | --- | --- |
| **Low** | High (n=11) | Obstructive | 8 | 15.7% |
|  |  | Non-obstructive | 3 | 5.9% |
|  | Low (n=40) | Obstructive | 4 | 7.8% |
|  |  | Non-obstructive | 36 | 70.6% |
| **Low Subtotal** |  |  | **51** | **100.0%** |
| **Medium** | High (n=49) | Obstructive | 36 | 18.6% |
|  |  | Non-obstructive | 13 | 6.7% |
|  | Low (n=145) | Obstructive | 27 | 13.9% |
|  |  | Non-obstructive | 118 | 60.8% |
| **Medium Subtotal** |  |  | **194** | **100.0%** |
| **High** | High (n=67) | Obstructive | 63 | 48.1% |
|  |  | Non-obstructive | 4 | 3.1% |
|  | Low (n=64) | Obstructive | 33 | 25.2% |
|  |  | Non-obstructive | 31 | 23.7% |
| **High Subtotal** |  |  | **131** | **100.0%** |

**Supplemental Table 3. Complete Reclassification pathways from HEART Score to angiographic outcome for “rule-out” performance of Infrasensor**

| **HEART**  **Score** | **Infrasensor** | **Severe Coronary Obstruction** | **N** | **% of HEART Category** |
| --- | --- | --- | --- | --- |
| **Low**  **(≤3)** | High (n=44) | Obstructive | 23 | 35.9% |
|  |  | Non-obstructive | 21 | 32.8% |
|  | Low (n=20) | Obstructive | 2 | 3.1% |
|  |  | Non-obstructive | 18 | 28.1% |
| **Low Subtotal** |  |  | **64** | **100.0%** |
| **Medium**  **(4-6)** | High (n=177) | Obstructive | 123 | 50.6% |
|  |  | Non-obstructive | 54 | 22.2% |
|  | Low (n=66) | Obstructive | 0 | 0.0% |
|  |  | Non-obstructive | 66 | 27.2% |
| **Medium Subtotal** |  |  | **243** | **100.0%** |
| **High**  **(≥7)** | High (n=156) | Obstructive | 145 | 86.8% |
|  |  | Non-obstructive | 11 | 6.6% |
|  | Low (n=11) | Obstructive | 3 | 1.8% |
|  |  | Non-obstructive | 8 | 4.8% |
| **High Subtotal** |  |  | **167** | **100.0%** |
