## Supplementary material for "A Wearable Infrared Sensor for Detecting Non-ST Segment Elevation Acute Coronary Syndromes": Suplemental Material Checklist

TRIPOD+AI Checklist

| **Section/Topic** | **Item** | **D/E** | **Checklist Item** | **Information from Paper** |
| --- | --- | --- | --- | --- |
| **TITLE** | | | | |
| Title | 1 | D;E | Identify study as developing/evaluating prediction model, target population, outcome | Development and external validation of ML model using Infrasensor for detecting high-grade obstructive NSTE-ACS including OMI in patients with suspected acute coronary syndrome |
| **ABSTRACT** | | | | |
| Abstract | 2 | D;E | See TRIPOD+AI for Abstracts checklist | Abstract describes multicenter study (n=595), leave-one-cohort-out validation, AUC 0.87 for rule-in, AUC 0.89 for rule-out |
| **INTRODUCTION** | | | | |
| Background | 3a | D;E | Explain healthcare context and rationale | Diagnostic context: NSTE-ACS detection. Rationale: Current diagnosis relies on serial troponin and ECG taking 6-12 hours; OMI missed in 60% using standard ECG criteria; need for rapid point-of-care tool |
|  | 3b | D;E | Describe target population and intended purpose | Target: Patients presenting with suspected NSTE-ACS. Purpose: Early risk stratification and triage - rule-in high-grade obstruction for urgent angiography, rule-out low-risk patients. Users: Emergency physicians, cardiologists |
|  | 3c | D;E | Describe known health inequalities | Rural/underserved areas face delayed diagnoses; Discussion addresses potential to improve care in resource-limited settings |
| Objectives | 4 | D;E | Specify study objectives | Primary: Evaluate Infrasensor for detecting severe coronary stenosis/OMI. Secondary: Identify patients without NSTE-ACS or adverse 30-day outcomes. Study includes both development and external validation |
| **METHODS** | | | | |
| Data | 5a | D;E | Describe data sources | Prospective multicenter cohort study. Development: Data from 5 geographic cohorts (4 India, 1 US). Validation: Leave-one-cohort-out external geographic validation. 200 healthy controls from US for calibration |
|  | 5b | D;E | Specify dates of data collection | Enrollment dates mentioned as 'Date through Date' (placeholder in manuscript). 30-day follow-up for outcomes |
| Participants | 6a | D;E | Specify study setting | 13 sites: 4 US sites (n=85) and 9 India sites (n=510). Settings include emergency departments and cardiac care facilities |
|  | 6b | D;E | Describe eligibility criteria | Inclusion: Adults ≥18 years with suspected ACS undergoing NSTEMI evaluation. Exclusions: STEMI, tattoos/scars at wrist, pulmonary HTN, implantable cardiac devices, pregnancy, malignancy, hemodynamic instability, altered consciousness |
|  | 6c | D;E | Give treatment details | Standard of care for ACS maintained. Revascularization at discretion of interventionalist. 42% PCI, 6% CABG among high-grade obstructive NSTE-ACS patients |
| Data preparation | 7 | D;E | Describe preprocessing and quality checking | Raw Infrasensor recordings underwent standardized signal-quality checks to exclude noisy segments. Features imputed (median within training folds) and scaled. Outlier detection performed. Same preprocessing across cohorts |
| Outcome | 8a | D;E | Define outcome and time horizon | Primary endpoint: ≥70% stenosis in epicardial coronary or ≥50% left main, assessed at coronary angiography during hospitalization. Secondary: Composite of NSTE-ACS diagnosis, revascularization, or death within 30 days |
|  | 8b | D;E | Describe outcome assessors | Core lab interventional cardiologist (blinded) reviewed all angiograms. Second blinded review for significant/severe stenosis. Majority consensus for discrepancies |
|  | 8c | D;E | Actions to blind outcome assessment | Core lab reviewer blinded to local site adjudication, clinical data, and Infrasensor output |
| Predictors | 9a | D | Describe choice of predictors | Features derived from Infrasensor time series: statistical (mean, SD, variance, skewness, kurtosis), percentiles, time-domain (RMS, energy, autocorrelation), frequency-domain (FFT, Welch PSD), complexity (Shannon entropy), time-frequency (STFT). Near-zero variance features excluded |
|  | 9b | D;E | Define all predictors | All predictors from 3-minute Infrasensor reading. Detailed in Supplementary Material Section I: statistical descriptors, percentiles, MAD, RMS, energy metrics, zero-crossing rate, FFT features, Welch PSD, Shannon entropy, STFT spectrograms, Hurst exponent |
|  | 9c | D;E | Describe predictor assessors | Infrasensor data collected by trained staff after ECG, before angiography. Automated signal acquisition (3-minute protocol) |
| Sample size | 10 | D;E | Explain study size | Total n=795 (595 NSTE-ACS patients + 200 healthy controls). Primary analysis n=469 (underwent angiography). Leave-one-cohort-out validation with 5 cohorts of approximately equal size for robust external validation |
| Missing data | 11 | D;E | Describe handling of missing data | Features with excessive missingness excluded. Remaining features imputed using median within training folds. Not all patients had FCU (n=351) or CMR (n=127) - analyzed as substudies |
| Analytical methods | 12a | D | Describe how data were used | Leave-one-cohort-out cross-validation: models trained on 4 cohorts, evaluated on held-out cohort. 200 healthy controls included in training for calibration. Separate models for rule-in (n=469 with angiography) and rule-out (n=595) |
|  | 12b | D | Describe predictor handling | Features computed from time series. Scaling applied. No transformation or functional form specification detailed beyond feature engineering |
|  | 12c | D | Specify model type, hyperparameter tuning, internal validation | Final model: 3-stage LightGBM ensemble. Stage 1: Feature selection LightGBM. Stage 2: Two calibrated LightGBM classifiers with different configurations. Stage 3: Probability averaging. Hyperparameters tuned via randomized search with early stopping. Full parameters in Supplemental Table 1 |
|  | 12d | D;E | Describe heterogeneity handling across clusters | Q-statistic heterogeneity test used to assess variation in model performance across cohorts. pHET <0.05 indicates significant heterogeneity. Internal-external cross-validation with pooled inference |
|  | 12e | D;E | Specify performance measures | AUC (ROC) with 95% CI via bootstrapping. Sensitivity, specificity, PPV, NPV at 0.5 threshold. DeLong test for AUC comparison. Sankey diagrams for reclassification. SHAP analysis for feature importance |
|  | 12f | E | Describe model updating |  |
|  | 12g | E | Describe how predictions calculated | Three-stage ensemble: (1) Feature selection, (2) Two calibrated LightGBM models with Platt scaling, (3) Average of probabilities. Threshold 0.5 for classification |
| Class imbalance | 13 | D;E | Describe class imbalance methods | Class weights applied (positive:negative ≈ 5:1) in gradient boosting models. Probability calibration via Platt scaling (sigmoid). 200 healthy controls added to enrich negative class for calibration |
| Fairness | 14 | D;E | Describe fairness approaches | Subgroup analyses by demographic and comorbidity characteristics showed no significant effect modification. Model performance consistent across clinically relevant subgroups (Table 2) |
| Model output | 15 | D | Specify model output | Probability output (0-1). Classification threshold: 0.5 (MAP decision rule). Rule-in: positive if probability ≥0.5. Rule-out: separate model for freedom from ACS |
| Training vs evaluation | 16 | D;E | Identify differences between dev and eval data | Geographic differences: India (Groups 1-4) vs US (Group 5). US patients older, higher BMI, more comorbidities. NSTEMI more common in US, unstable angina more common in India. Same eligibility criteria and outcome definitions |
| Ethical approval | 17 | D;E | Name IRB and consent process | Study approved by each institution's IRB. Adhered to Good Clinical Practices. Informed consent obtained from all participants |
| **OPEN SCIENCE** | | | | |
| Funding | 18a | D;E | Give funding source | Sponsored by RCE Technologies. PPS supported by NIH/NHLBI (1R01HL173998-01A1, 3U01HL088942-17S, 1P50MD017356-01) and NSF (#2125872) |
| Conflicts | 18b | D;E | Declare conflicts of interest | Provided on page 26 of main text |
| Protocol | 18c | D;E | Indicate protocol access | NA |
| Registration | 18d | D;E | Provide registration information | NA |
| Data sharing | 18e | D;E | Provide data availability details | The datasets generated during this study are not publicly available due to institutional regulations and ethics/IRB approval terms. De-identified data are available from the corresponding author upon reasonable request, subject to institutional approval and a data use agreement. |
| Code sharing | 18f | D;E | Provide code availability details | Custom code available from corresponding author upon reasonable request for academic research purposes |
| **PATIENT & PUBLIC INVOLVEMENT** | | | | |
| PPI | 19 | D;E | Provide PPI details | NA |
| **RESULTS** | | | | |
| Participants | 20a | D;E | Describe participant flow | 795 enrolled (595 NSTE-ACS + 200 controls). 469/595 (79%) underwent angiography. Outcomes: 215 (45.8%) high-grade obstruction, 91 (19.4%) OMI. 30-day: 11 ACS readmissions, 2 PCI, 3 CABG, 2 deaths |
|  | 20b | D;E | Report characteristics | Table 1 provides demographics by 5 groups. Median age 55-65 years. Male 56.7-78%. Diabetes 32.6-50%. Notable differences: US older, higher BMI, more comorbidities. Troponin patterns varied significantly |
|  | 20c | E | Compare dev vs eval distributions | Table 1 shows distribution across 5 cohorts. US vs India differences noted in age, BMI, comorbidities, clinical presentation |
| Model development | 21 | D;E | Specify participants in each analysis | Rule-in model: n=469 (with angiography), 215 events.  Rule-out model: n=595, 354 true positives. CMR substudy n=127, FCU substudy n=351 |
| Model specification | 22 | D | Provide full model details | Three-stage LightGBM ensemble.  Full hyperparameters in Supplemental Table   1. Stage 1: n_estimators=550, lr=0.05, Num leaves=32. 2. Stage 2 Model 1: n_estimators=300, lr=0.05. 3. Stage 2 Model 2: n_estimators=620, lr=0.04. 4. Calibration via Platt scaling. |
| Model performance | 23a | D;E | Report performance with CIs | Rule-in: AUC 0.87 (95% CI: 0.84-0.90), specificity 90%, PPV 84%. Rule-out: AUC 0.89 (95% CI: 0.87-0.92), sensitivity 99%, NPV 96%. Subgroup analyses showed consistent performance (Table 2) |
|  | 23b | D;E | Report heterogeneity across clusters | Q-statistic heterogeneity test performed. No significant effect modification by demographic or comorbidity characteristics |
| Model updating | 24 | E | Report updating results | Not applicable - no model updating performed |
| **DISCUSSION** | | | | |
| Interpretation | 25 | D;E | Give overall interpretation | Infrasensor outperform |
| Limitations | 26 | D;E | Discuss limitations | Hospital-based cohort only. Impact on hard outcomes not yet determined. Biological mechanism incompletely understood. False positive rate ~10% could lead to unnecessary procedures. Geographic/demographic differences between sites |
| Usability | 27a | D | Describe handling of poor input data | Signal quality checks exclude noisy segments. 45-second background calibration before measurement |
|  | 27b | D | Specify user expertise required | Minimal expertise: 70% alcohol wipe on wrist, strap device, 3-minute acquisition. Results transmitted to cloud platform automatically |
|  | 27c | D;E | Discuss future research | RCT needed for impact on outcomes. Pre-hospital/ambulance setting evaluation. Integration with AI-enhanced ECG. Cost-effectiveness analysis. Stable outpatient and post-PCI monitoring applications |

**Notes:**

*D = items relevant to development; E = items relevant to evaluation; D;E = both*

*Green cells contain information extracted from the paper. Items not explicitly addressed are noted.*
